# Ward-Level Factors Associated with Methicillin-Resistant *Staphylococcus aureus* Acquisition – an Electronic Medical Records study in Singapore

**DOI:** 10.1101/2020.07.07.20148155

**Authors:** Zaw Myo Tun, Dale A. Fisher, Sharon Salmon, Clarence C. Tam

**Author notes:** Corresponding author: (ZMT).

## Abstract

**Background:** Methicillin-Resistant *Staphylococcus aureus* (MRSA) is endemic in hospitals worldwide. When patients are transferred between wards within a hospital, their risk of acquiring MRSA may change. In this study, we investigated how ward characteristics and connectivity are associated with MRSA acquisition.

**Methods:** We analysed electronic medical records on patient transfers and MRSA screening of in-patients at an acute-care tertiary hospital in Singapore to investigate whether ward characteristics and connectivity within the hospital network were associated with MRSA acquisition rates over a period of four years.

**Results:** Most patient transfers concentrated in a stable core network of wards. Factors associated with increased rate of MRSA acquisition were ward MRSA admission prevalence (rate ratio (RR): 1.50, 95% CI: 1.28, 1.71, per one percentage point increase), admission to a critical care ward (RR: 1.86, 95% CI: 1.14, 3.06) and average number of patients in the ward on a typical day (RR: 1.31, 95% CI: 1.02, 1.68, for every 10 patients quarterly). Admission to an oncology ward (RR: 0.61, 95% CI: 0.40, 0.93) (compared to medical ward), and median length of stay (RR: 0.71, 95% CI: 0.54, 0.93) were associated with lower acquisition rates. We did not find evidence that network measures of ward connectivity, including in-degree, weighted in-degree, influenced MRSA acquisition rate after adjusting for other ward characteristics.

**Conclusion:** Ward MRSA admission prevalence, critical care ward, ward patient capacity, ward specialty, and median length of stay, rather than relative connectivity of the ward in the hospital network were associated with MRSA acquisition.

## Introduction

Since its emergence in the 1960s, Meticillin-Resistant *Staphylococcus aureus* (MRSA) has become endemic in hospitals worldwide, causing significant health and financial burden (1, 2), accounting for >20% of *Staphylococcus aureus* bloodstream infections globally (3). In high-income settings, the incidence of hospital-onset MRSA infection has declined over time, although progress in controlling MRSA has plateaued in recent years (4-6).

In Singapore, a high-income city state in Asia, acute care public hospitals initiated a multi-pronged MRSA control strategy from 2006 (7-9). This strategy, comprising active surveillance screening for MRSA, hand hygiene promotion and compliance auditing, cohorting of colonized patients with contact precaution, and other measures, led to a substantial reduction in hospital-acquired MRSA bacteremias (7). Despite this, MRSA remains endemic in healthcare settings. A point prevalence survey in 2014 indicated that 11.8% of patients in a large tertiary public hospital were colonized by MRSA. The prevalence was higher in intermediate (29.9%) and long-term (20.4%) care facilities (10).

Colonization with MRSA is a major risk factor for invasive disease. Numerous factors are known to be associated with MRSA colonization, including patient characteristics such as older age (11), previous hospitalization (11, 12), intensive care unit (ICU) admission (13), chronic illness (14), exposure to other patients known to be colonized with MRSA (15-17), antibiotic use (12, 15), prolonged hospital stay (16, 18), and receiving medical procedures during hospitalization (12). Other factors include colonization pressure (13, 16, 19), infection control practices (such as hand hygiene compliance (7), cohorting (7), environmental decontamination (20), MRSA colonization status of healthcare staff (21)), and organizational factors (such as staff to patient ratio (22), bed occupancy rate (23), patient capacity of a ward (24)).

Variation in infection control practices and organizational factors between hospital functional units means that MRSA acquisition risk experienced by a patient is likely to change if they are transferred between wards within a hospital. Studies investigating MRSA acquisition risks associated with intra-hospital patient transfer are rare, because of the need for longitudinal data on patient transfers and MRSA colonization status with high temporal resolution. A case-control study by Dziekan and colleagues found a linear relationship: the greater the number of between-ward transfers, the higher the risk of MRSA acquisition (25). This led us to assess whether greater ward connectivity in terms of patient transfer influences MRSA acquisition risk. In this study, we used high-resolution electronic medical records of in-patient ward transfers from a large public acute care hospital in Singapore, together with active MRSA admission screening data, to identify ward characteristics associated with MRSA acquisition.

## Methods

### Data sources

We used a dataset of in-patient electronic medical records from the National University Hospital (NUH), Singapore spanning January 1, 2010 to December 31, 2013 (the Patient Affordability Simulation System (PASS)). In addition, we obtained data on in-hospital MRSA acquisitions over the same period from the hospital’s MRSA active surveillance culture data set, as well as ward-level hand hygiene compliance from audit data.

### Patient Affordability Simulation System (PASS)

PASS records hospital service use and cost information (26). The following variables were available in the dataset: ward number, ward specialty, patients’ age, and timestamps for patients’ admission, transfers between wards and discharge.

### MRSA Active Surveillance Cultures

Active MRSA screening is implemented in 36 out of 64 in-patient wards. Other low-risk wards do not routinely perform active screening and include obstetric, pediatric, psychiatric and acute stay wards. The screening process involves obtaining nasal, axillary, and groin (NAG) swabs at admission, transfer, and discharge. These samples are cultured on selective chromogenic agar. Swabs are obtained on the day of or one day before/after the admission or transfer, and on the day of or one day before discharge. The exceptions are patients hospitalized for <48 hours, those with a MRSA positive result in a previous hospitalization, and deceased patients. MRSA results from clinical isolates are not captured in the active screening database.

### Hand-hygiene compliance

Infection control liaison nurses perform random audits in 40 in-patient wards once a month. Twenty observations of healthcare staff hand hygiene activities are recorded clandestinely at any time of the day (27). Hand hygiene compliance is defined as per WHO guidance: the number of hand hygiene activities performed as a percentage of the total number of hand hygiene opportunities (28). The data were available quarterly.

### Data linkage

A third-party analyst who was not a study team member linked these data sets using unique patient identifiers and anonymized them before providing access to the study team.

### Network analysis

We constructed a weighted directed network using patient transfer data to understand how hospital wards are connected. The network comprised all 64 in-patient wards represented as nodes. Ward connectivity through patient transfers was represented as directed edges linking the origin and destination wards. Edges were weighted based on the number of patients transferred over a specific period.

To investigate the association of ward connectivity and MRSA acquisition rates, we used in-degree and weighted in-degree as network centrality measures. The former represents the number of other wards from which a focal ward receives at least one patient, while the latter reflects the number of patients a focal ward receives from other wards. We constructed 16 quarterly networks from patient transfer data and computed quarterly network measures for consistency with the temporal resolution of hand hygiene compliance data.

### Inclusion and exclusion criteria

We included in the analysis in-patient admissions to one of the 36 active screening wards lasting >48 hours. We defined a hospitalization episode as the period between admission to and discharge from the hospital. One hospitalization episode could contain one or more spells, defined as the period from entry into to exit from a hospital ward.

We excluded from analysis episodes with a positive or no screening result at admission; episodes of patients younger than 15 (pediatric patients are not routinely screened for MRSA); and episodes with a negative MRSA result at admission but no subsequent MRSA screening results.

A MRSA acquisition event was defined as an initially MRSA-negative patient who was found positive during a hospitalization episode. For each ward, we estimated MRSA acquisition rate, defined as the number of MRSA acquisitions per 100 patient-weeks. We computed patient-weeks at risk for each ward by summing the total time spent by patients in that ward. For patients who acquired MRSA, their contribution to patient-weeks at risk was censored at the time of collecting a sample positive for MRSA.

### Statistical analysis

We used mixed-effects Poisson regression to identify ward-level factors associated with MRSA acquisition. The outcome was the total number of MRSA acquisitions in a specific ward in each quarter. The natural logarithm of the total patient-weeks at risk was used as an offset. We modelled wards as a random intercept and time (in quarters) as a random slope to account for ward-level variability in MRSA acquisition rates and their trends, respectively.

We extracted nine explanatory variables from the available data sources. Time-varying variables included quarterly ward in-degree and weighted in-degree, number of patients in a ward on a typical day, ward MRSA admission prevalence, length of stay, and hand hygiene compliance. Time-invariant variables were critical care ward (i.e., ICU and high dependency unit (HDU)), ward specialty (medical, surgical, orthopedics, oncology, and other), and presence of cohorting beds for MRSA-positive patients. The number of patients in a ward on a typical day was the quarterly average number of patients registered on the 15_th_ of each month. This was considered a proxy for a ward’s patient capacity.

We considered ward MRSA admission prevalence as a proxy for colonization pressure. In a hospital ward, the colonization pressure of a pathogen is mainly influenced by admission of patients who are already colonized (29). Admission prevalence was measured using only the first hospitalization of all patients between 2010 and 2013. This is because, as per the screening protocol, no screening is performed in subsequent hospitalizations if a patient has been previously identified as MRSA positive; including all hospitalization episodes would underestimate the admission prevalence.

### Sensitivity Analyses

Hand hygiene compliance audit was not implemented in some of the active MRSA screening wards and thus data were unavailable. Consequently, we could not include these wards in modelling hand hygiene compliance data. We therefore compared the results of the multivariable models with and without this variable.

In 299 out of 2,370 (12.8%) MRSA-positive hospitalization episodes, information on MRSA screening was missing in at least one ward spell prior to that in which the positive result was obtained. For these episodes, we could not determine the exact ward in which patients acquired MRSA. To assess the impact of missing MRSA screening information on the results, we conducted sensitivity analyses using five scenarios: (1) complete case analysis – we only included episodes with complete screening results for all spells; (2) mid-point analysis – we assumed that MRSA acquisition occurred in the ward the patient was in at the mid-point between the last known negative result and the positive result; in the next three scenarios, we probabilistically attributed MRSA acquisition to spells with missing MRSA results by random selection (3) using equal probabilities; (4) using a probability weighted by the patient’s length of stay in each spell (16, 18, 19, 30, 31); and (5) using a probability weighted by both length of stay and MRSA admission prevalence in each spell (16, 19, 30, 31). For scenarios 1 and 2, we obtained point estimates and confidence intervals from the multivariable model. For scenarios 3 to 5, we iterated the imputation and model fitting 10,000 times to obtain an empirical distribution of estimates for each parameter. We took the median, 2.5_th_ and 97.5_th_ percentiles of these distributions as the point estimate, and lower and upper confidence bounds, respectively.

We considered scenario five as the main analysis as we deemed its assumptions to be more realistically capture the uncertainty associated with missing screening data. Analyses were carried out using R (version 3.5.2) (32), igraph package (33), and lme4 package (34).

### Ethics review

Ethical exemption for this secondary data analysis was obtained from the National Healthcare Group Domain Specific Review Board (reference number: 2018/00890).

## Results

We successfully linked 92% of MRSA screening results in the MRSA Active Surveillance Cultures dataset to PASS. A total of 64,362 hospitalization episodes were eligible to investigate factors associated with MRSA acquisition (Fig 1).

**Fig 1.**
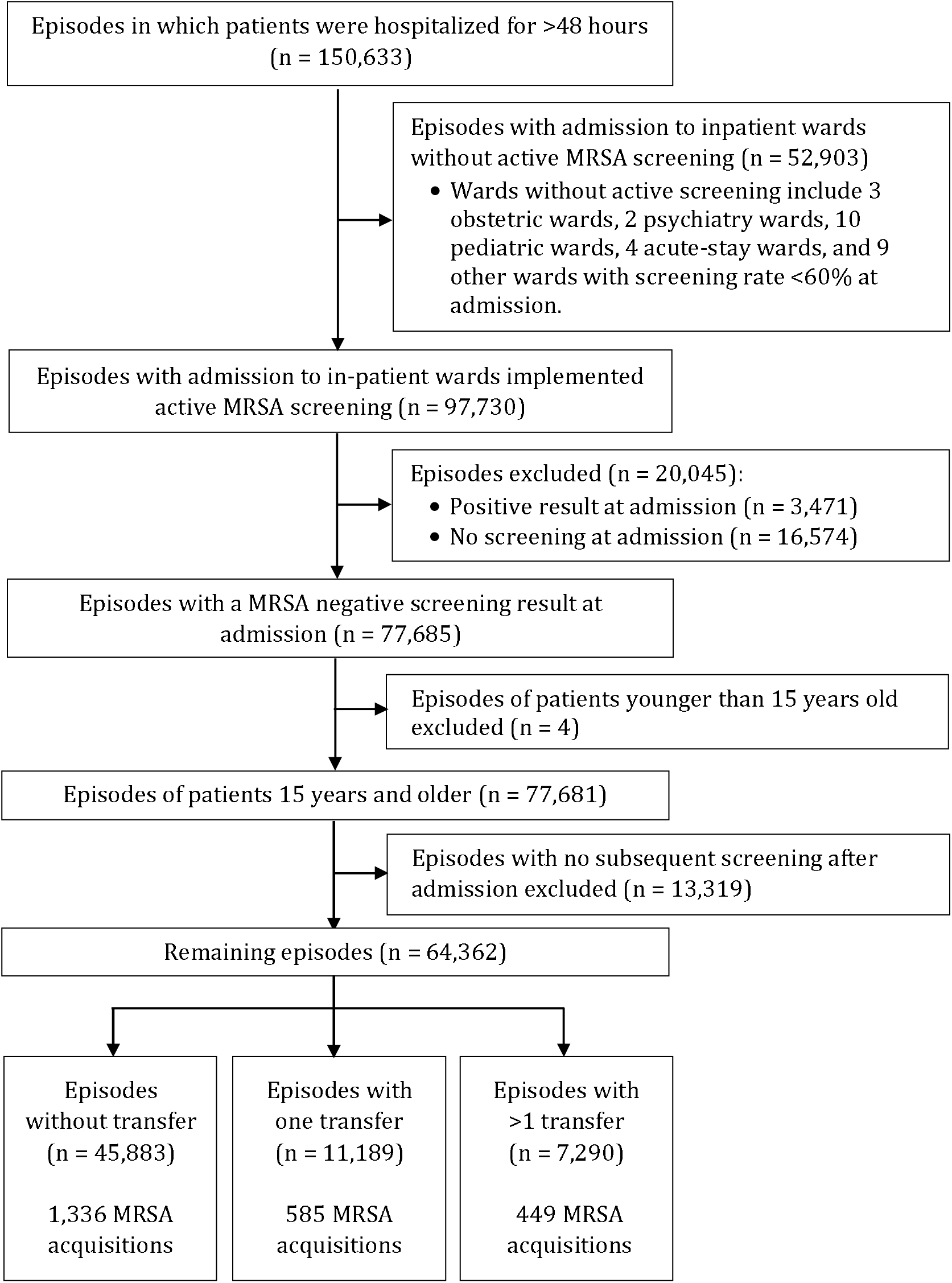
Hospitalization episodes from 2010 to 2013 in National University Hospital, Singapore included in the analysis.

### Characteristics of in-patient wards

In quarterly networks of 64 in-patient wards, in-degree was highest in Ward 1 Surgery (HDU), Ward 1 Medical (ICU/HDU), and Ward 1 Isolation in most quarters over four years. Weighted in-degree was highest in Ward 1 Surgery (HDU), Ward 3 Cardiac, and Ward 4 Surgery. On the other hand, we observed lowest values in both in-degree and weighted in-degree in Ward 2 Psychiatry, Ward 2 Other, and Ward 5 Coronary care/Cardiac medical. Characteristics of all 64 in-patient wards are further enumerated in (S4 Table).

Of 36 active screening wards, 8 (22%) were critical care wards; 8 (22%) contained MRSA-cohorting beds. Median MRSA admission prevalence was 1.8% (range: 0, 4.7); median hand hygiene compliance was 70.4% (range: 61.8, 83.7). Median in-degree and weighted in-degree were 21.5 (range: 6, 30) and 137 (range: 16, 410.5), respectively (Table 1).

**Table 1.** Characteristics of wards with MRSA active screening at the National University Hospital, Singapore in 2010-2013.

| Time invariant variable | No. wards | % |
| --- | --- | --- |
| Critical care wards | 8 | 22 |
| Presence of MRSA cohorting beds | 8 | 22 |
| Ward specialty |  |  |
| Medical | 7 | 19 |
| Surgical | 10 | 28 |
| Oncology | 8 | 22 |
| Orthopedics | 3 | 8 |
| Other | 8 | 22 |
| Time varying variable* | Median | Range |
| Number of patients in a ward on a typical day | 21.8 | 2.7, 54.2 |
| Length of stay (days) | 3.3 | 2.6, 7.7 |
| Admission prevalence of MRSA | 1.8 | 0, 4.7 |
| Hand hygiene compliance (%) | 70.4 | 61.8, 83.7 |
| Measures of ward connectivity |  |  |
| In-degree | 21.5 | 6, 30 |
| Weighted in-degree | 137 | 16, 410.5 |

MRSA, Methicillin-resistant *Staphylococcus aureus*
\* Median of quarterly measurements was quantified for each ward. Median and range of these values from 36 in-patient wards with active MRSA screening were presented.

### MRSA acquisition rates

MRSA acquisitions were identified in 2,370 of 64,362 (3.7%) hospitalization episodes over four years. In the main analysis, the median overall acquisition rate was 3.3 acquisitions per 100 patient-weeks (95% confidence interval (CI): 3.2, 3.5). The impact of missing MRSA results in spells prior to that in which the positive result was obtained was small: the maximum range of variability in 16 quarters over 10,000 iterations was only 0.2 acquisitions per 100 patient-weeks (S1 Fig). The acquisition rates were higher in the hospital wards of the following specialties: surgery, geriatric medicine, orthopedics, and cardiac. Overall MRSA acquisition rates by ward are shown in S2 Table.

### Factors associated with ward-level MRSA acquisition rates

In our main analysis, factors associated with a higher MRSA acquisition rate were: ward admission prevalence (rate ratio (RR): 1.50, 95% CI: 1.31, 1.72, per one percentage point increase), critical care ward (RR: 1.86, 95% CI: 1.14, 3.06) and average number of patients on a typical day (RR: 1.31, 95% CI: 1.02, 1.68, for every 10 additional patients). On the other hand, oncology ward (RR: 0.61, 95% CI: 0.40, 0.93) (compared to medical ward), and median length of stay (RR: 0.71, 95% CI: 0.54, 0.93) were associated with a lower acquisition rate (Table 2). Sensitivity analyses showed that the direction of association was largely consistent across all scenarios (Fig 2).

**Table 2.** Ward characteristics associated with MRSA acquisition.

| Ward characteristics | Unadjusted RR (95% CI) | Adjusted RR (95% CI) |
| --- | --- | --- |
| Critical care wards |  |  |
| No | 1 | 1 |
| Yes | 1.09 (0.68, 1.76) | 1.86 (1.14, 3.06) |
| Presence of MRSA cohorting beds |  |  |
| No | 1 | 1 |
| Yes | 1.41 (0.94, 2.11) | 1.38 (0.95, 2.01) |
| Ward specialty |  |  |
| Medical | 1 | 1 |
| Surgical | 1.08 (0.70, 1.65) | 0.90 (0.62, 1.30) |
| Oncology | 0.43 (0.27, 0.70) | 0.61 (0.40, 0.93) |
| Orthopedics | 1.24 (0.66, 2.31) | 0.93 (0.56, 1.54) |
| Other | 1.10 (0.73, 1.66) | 1.20 (0.78, 1.84) |
| Number of patients in a ward on a typical day* (for each 10 additional patients) | 1.37 (1.09, 1.73) | 1.31 (1.02, 1.68) |
| Median length of stay (for each day increase) | 0.75 (0.59, 0.95) | 0.71 (0.54, 0.93) |
| Admission prevalence of MRSA (per one percentage point increase) | 1.50 (1.30, 1.72) | 1.50 (1.31, 1.72) |
| In-degree (for each additional ward) | 1.40 (0.76, 2.58) | 0.96 (0.51, 1.79) |
| Weighted in-degree (for each 10 additional patients) | 4.51 (1.21, 16.81) | 1.25 (0.32, 4.88) |

2 CI, Confidence Interval; RR, Rate Ratio

**Fig 2.**
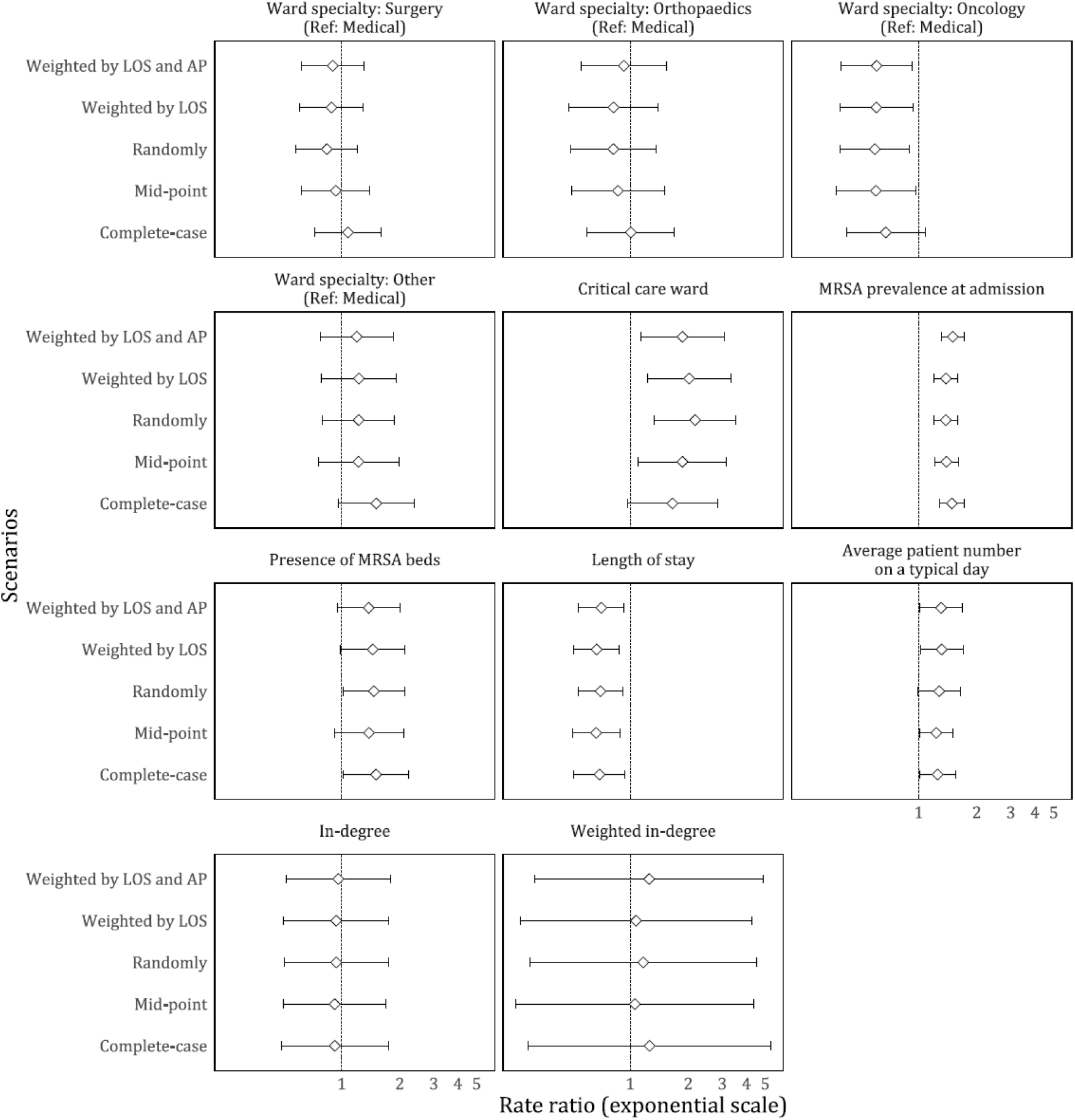
Sensitivity analyses accounting for the impact of spells without screening prior to a positive spell. In our main analysis, these spells were assigned MRSA acquisition with a probability weighted by LOS and AP of these spells. Each panel describes rate ratio with corresponding 95% confidence interval of each term in the multivariable models. AP, Admission prevalence; LOS, Length of stay

We compared the above model with one including hand hygiene compliance. The results of the latter showed that hand hygiene compliance itself was not associated with MRSA acquisition rate in the subset of wards for which this information was available, after controlling for other factors. However, compared to the main analysis, the estimates were substantially different for ward specialty, presence of MRSA cohorting beds, and median length of stay (S3 Table).

## Discussion

We used electronic medical records with high temporal resolution to understand in-patient ward connectivity in a large acute care hospital and ward characteristics associated with MRSA acquisition. We found that ward specialty, median length of stay, MRSA admission prevalence, critical care ward, average number of patients on a typical day were associated with MRSA acquisition rates. However, network measures of ward connectivity, such as the number of wards from which patients were received, or the number of patients received from other wards, were not associated with MRSA acquisition rates in wards.

MRSA acquisition rate was generally lower in oncology wards compared to other wards, after adjusting for potential confounders. The acquisition rate of a ward reflects a balance between the ward’s case mix (35) and how stringently infection control measures are implemented (36). Oncology wards, which tend to have patients at higher risk of infections, are likely to have stricter adherence to infection control measures, and our findings also suggest that improvements in infection control should be possible for other ward types.

We did not find evidence that ward connectivity measures (in-degree and weighted in-degree) influenced ward-level MRSA acquisition after adjusting for other ward characteristics. It should be noted that in our ward-level analysis, the connectivity measures used can only account for the total number of transfers between a ward pair, rather than the number of transfers for individual patients (25). For instance, a highly connected ward may have lower acquisition rate, perhaps because of better infection control measures, but it is possible that individual patients from this ward who undergo more transfers still experience higher MRSA acquisition risk.

The median hand hygiene compliance in NUH was 70.4%, comparable to large tertiary hospitals in Hong Kong (37) and Taiwan (38) using similar monitoring protocols. Differences in the results of multivariable models with and without hand hygiene compliance suggest that wards with and without the hand hygiene compliance data available differed with respect to the other characteristics investigated.

Although length of hospital stay is an important patient-level risk factor (16, 18, 19, 30, 31), our ward-level analysis showed the opposite: median length of stay was associated with a lower MRSA acquisition rate. This may reflect stricter infection control measures in the in-patient wards in which patient length of stay is longer.

In line with previous studies (13, 35, 39), we found that critical care ward status was associated with higher rate of MRSA acquisition. Critical care patients are known to be at particularly high risk for nosocomial infections, pointing to a need for more stringent infection control measures in these wards.

Several limitations should be considered when interpreting our findings. Firstly, the unavailability of MRSA results from clinical isolates means that we could not include a subset of MRSA acquisitions that are not identified through routine screening. However, the incidence of MRSA infections in NUH is <1 per 100 patient-weeks (40), so the impact of this is likely small. Secondly, we could not adjust for ward staffing level (22, 41), or MRSA colonization status and compliance with contact precaution measures of healthcare staff as this information is not routinely available (21, 42). Lastly, this ward-level analysis cannot account for individual-level differences in MRSA acquisition risk, including age, gender, comorbidities, and use of out-patient services. More detailed individual-level analyses could investigate the interaction between individual and ward-level risk factors. Nonetheless, electronic medical records provided objective measures of patients’ transfers through hospital that do not rely on recall and self-report, a major strength of this analysis.

Our analysis demonstrates an efficient use of linked electronic medical records and infection control data to comprehensively study the complexity of intra-hospital patient transfer patterns. Although we did not find evidence of the association of ward connectivity and MRSA acquisition, our findings highlighted other ward characteristics associated with MRSA acquisition rate, pointing to a need for further investigation and strengthening MRSA control efforts. Similar methods could be used to understand the transmission dynamics of other nosocomial organisms.

## Data Availability

We do not have the permission to share the data.

## Acknowledgements

The authors thank Mark Salloway and Joanne Chee on their assistance in data retrieval and linkage.

## Supporting Information

**S1 Fig.**
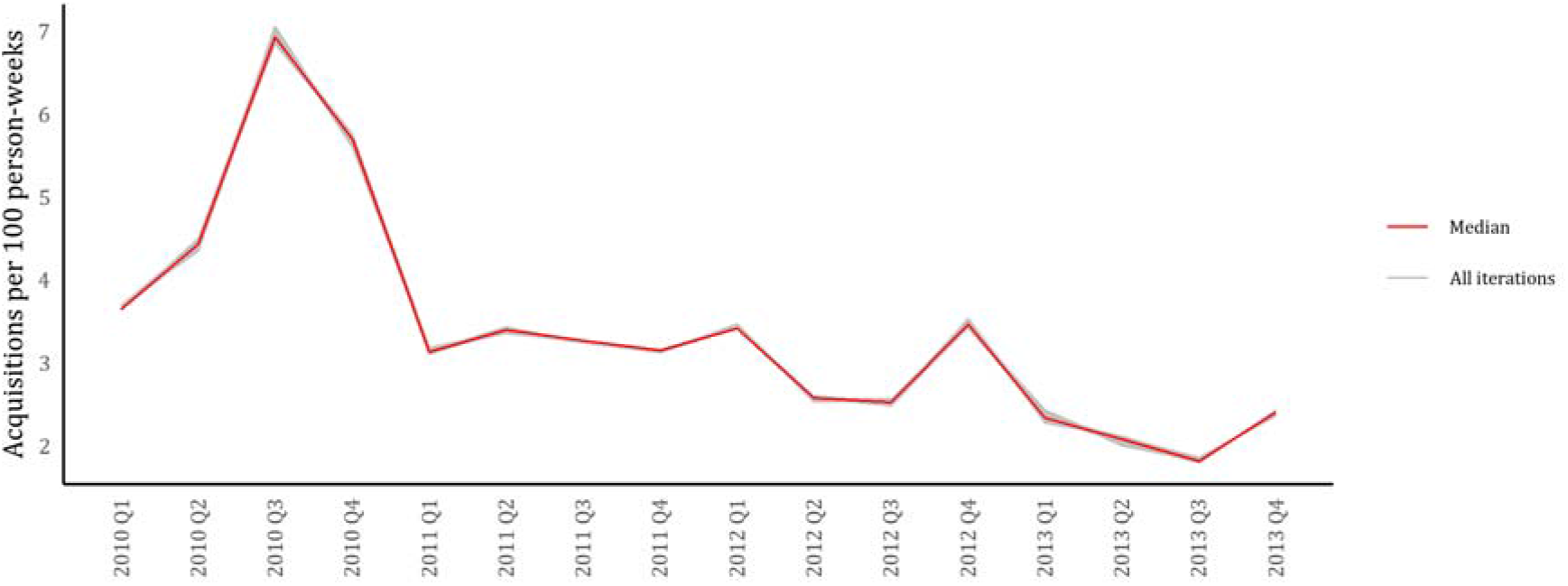
Quarterly MRSA acquisition rate among MRSA active screening wards at National University Hospital, 2010-2013.

**S2 Table.** MRSA acquisition rate in MRSA active screening wards of National University Hospital, 2010-2013.

| Ward<sup>^</sup> | Acquisition rate* | 95% CI | Median MRSA acquisitions<sup>#</sup> | Median person-week at risk<sup>#</sup> |
| --- | --- | --- | --- | --- |
| 11 Surgery, HDU | 12.9 | 4.2, 30.0 | 5 | 38.9 |
| 6 Medical, Geriatric Medicine | 5.8 | 5.0, 6.6 | 205 | 3539.0 |
| 2 Orthopaedics | 5.1 | 4.4, 6.0 | 163 | 3169.8 |
| 3 Cardiac | 4.8 | 4.2, 5.4 | 249 | 5194.4 |
| 4 Cardiac | 4.5 | 3.8, 5.2 | 173 | 3882.2 |
| 4 Surgery | 4.3 | 3.6, 5.0 | 161 | 3770.6 |
| 2 Cardio-Thoracic ICU | 4.2 | 3.1, 5.5 | 52 | 1241.6 |
| 2 Medical | 4.2 | 3.6, 4.8 | 173 | 4157.3 |
| 8 Medical, Neuro | 4.2 | 3.3, 5.2 | 72 | 1730.7 |
| 5 Medical, Renal | 3.9 | 3.3, 4.5 | 156 | 4038.0 |
| 5 Surgery | 3.8 | 3.2, 4.4 | 162 | 4311.7 |
| 1 Surgery, HDU | 3.5 | 2.7, 4.5 | 63 | 1804.1 |
| 2 Neurosurgery, HDU | 3.3 | 2.3, 4.7 | 31 | 931.2 |
| 1 Medical, ICU, HDU | 3.3 | 2.5, 4.4 | 54 | 1614.6 |
| 1 Cardiac | 3.2 | 2.2, 4.5 | 33 | 1037.9 |
| 3 Other ward | 3.0 | 2.1, 4.0 | 2 | 67.4 |
| 3 Surgery, ICU | 3.0 | 0.4, 10.7 | 25 | 864.3 |
| 1 Oncology, Medical | 2.9 | 1.9, 4.3 | 43 | 1456.9 |
| 6 Surgery | 2.9 | 2.2, 3.6 | 71 | 2571.1 |
| 2 Oncology | 2.8 | 2.2, 3.5 | 70 | 2454.7 |
| 4 Oncology | 2.5 | 1.9, 3.2 | 61 | 2465.4 |
| 2 Isolation | 2.5 | 1.9, 3.4 | 48 | 1890.3 |
| 5 Coronary Care, Cardiac Medical | 2.4 | 1.2, 4.1 | 12 | 509.1 |
| 9 Surgery | 2.2 | 1.4, 3.3 | 24 | 1084.1 |
| Medical, Surgery mixed | 2.1 | 1.5, 2.9 | 38 | 1796.4 |
| 3 Oncology | 1.8 | 1.3, 2.4 | 51 | 2849.0 |
| 1 Orthopaedics | 1.7 | 1.3, 2.2 | 59 | 3520.5 |
| 4 Medical, HDU | 1.7 | 0.6, 3.6 | 6 | 361.9 |
| 9 Medical | 1.4 | 0.8, 2.3 | 14 | 1011.8 |
| 8 Surgery | 1.0 | 0.5, 1.9 | 10 | 955.1 |
| 3 Orthopaedics | 0.8 | 0.3, 1.9 | 5 | 614.3 |
| 6 Oncology, Medical | 0.8 | 0.3, 1.7 | 7 | 862.7 |
| 10 Surgery | 0.6 | 0.1, 1.6 | 3 | 541.9 |
| 8 Oncology | 0.5 | 0.2, 1.2 | 5 | 949.8 |
| 7 Oncology | 0.5 | 0.2, 1.1 | 5 | 1023.5 |
| 5 Oncology, HDU | 0.2 | 0, 0.7 | 2 | 1068.2 |

CI, Confidence interval; HDU, High dependency unit; ICU, Intensive care unit
\* Number of MRSA acquisitions per 100 person-weeks
<sup>^</sup> The ward numbers are masked

**S3 Table.** Multivariable analysis with and without hand hygiene compliance.

| Ward characteristics | Model including hand hygiene compliance<br>Adjusted RR (95% CI) | Final model<br>Adjusted RR (95% CI) |
| --- | --- | --- |
| Critical care wards |  |  |
| No | 1 | 1 |
| Yes | 1.83 (1.20, 2.79) | 1.86 (1.14, 3.06) |
| Presence of MRSA cohorting beds |  |  |
| No | 1 | 1 |
| Yes | 1.78 (1.42, 2.22) | 1.38 (0.95, 2.01) |
| Ward specialty |  |  |
| Medical | 1 | 1 |
| Surgical | 1.02 (0.80, 1.28) | 0.90 (0.62, 1.30) |
| Oncology | 1.09 (0.81, 1.48) | 0.61 (0.40, 0.93) |
| Orthopedics | 1.16 (0.83, 1.61) | 0.93 (0.56, 1.54) |
| Other | 1.67 (1.26, 2.20) | 1.20 (0.78, 1.84) |
| Number of patients in a ward on a typical day* (for each 10 additional patients) | 1.57 (1.22, 2.00) | 1.31 (1.02, 1.68) |
| Median length of stay (for each day increase) | 1.02 (0.77, 1.36) | 0.71 (0.54, 0.93) |
| Admission prevalence of MRSA (per one percentage point increase) | 1.48 (1.28, 1.70) | 1.50 (1.31, 1.72) |
| In-degree (for each additional ward) | 1.12 (0.61, 2.04) | 0.96 (0.51, 1.79) |
| Weighted in-degree (for each 10 additional patients) | 1.50 (0.57, 3.97) | 1.25 (0.32, 4.88) |
| Hand hygiene compliance (for each percentage point increase) | 0.96 (0.90, 1.02) |  |

CI, Confidence Interval; RR, Rate Ratio

**S4 Table.**
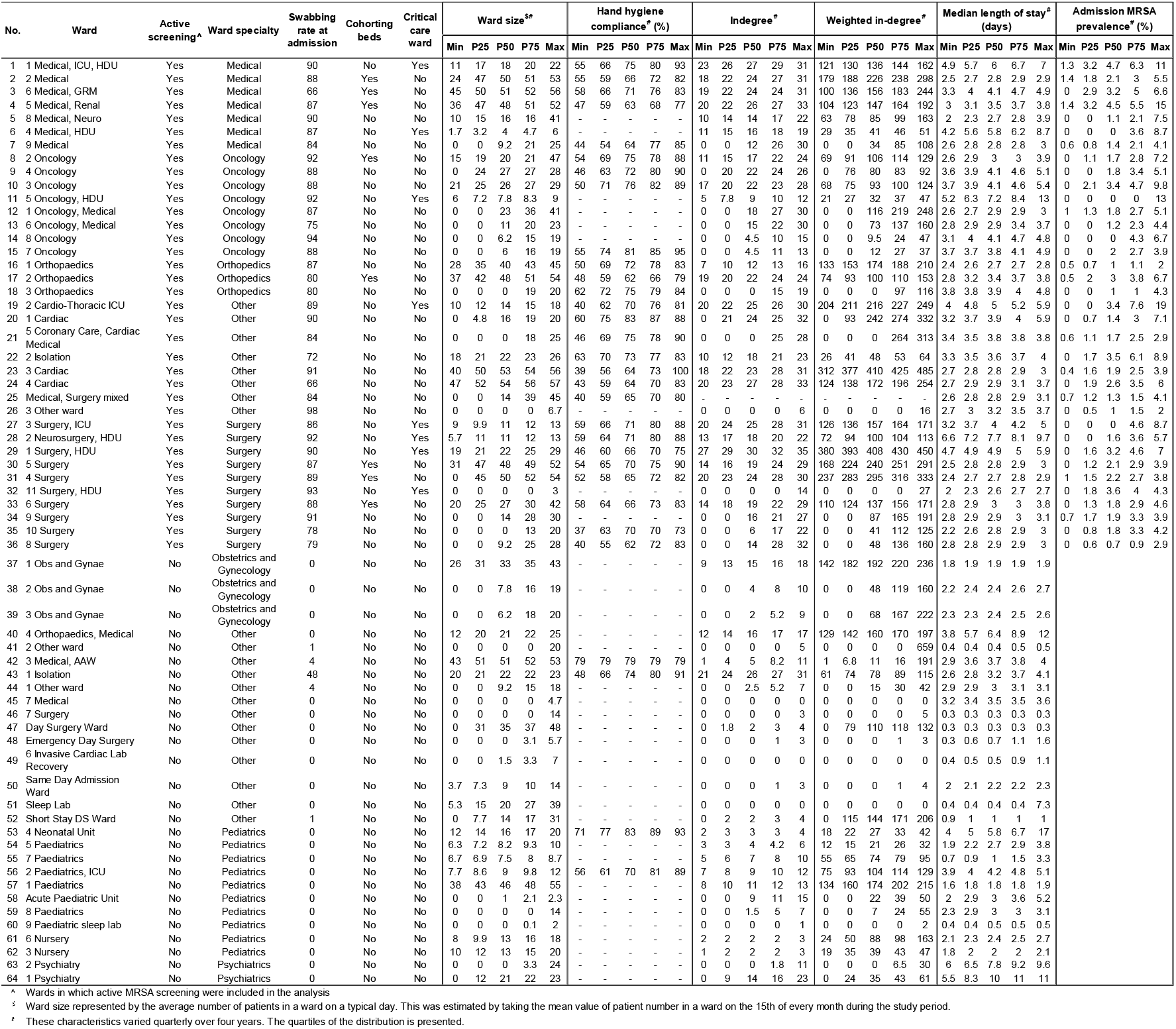
Characteristics of 64 in-patient wards of National University Hospital.

<This table is included separately in a excel workbook (sheet name: ward characteristics) in the submission.>

## Notes

### Competing Interest Statement

The authors have declared no competing interest.

### Funding Statement

No funding was received for this work.

